# Geospatial Patterns of Cariogenic Feeding Practices Among Children Aged 6-23 Months in Ghana: A Spatial and Multilevel Analysis of the 2022 Ghana Demographic and Health Survey

**DOI:** 10.64898/2026.08.06.26359897

**Authors:** Osman Abdul-Fatawu Iddrisu, Adiza Isma-il, Abubakar Hudu Siddick, Abubakar Iddrisu Siddiq

## Abstract

**Background:** Early childhood caries remains one of the most prevalent, and among the most neglected, chronic diseases of childhood in low- and middle-income countries, and cariogenic feeding practices introduced in infancy are a principal, modifiable driver. National, spatially explicit estimates of such practices are lacking for Ghana, constraining geographically targeted interventions.

**Methods:** We analysed 3,007 children aged 6-23 months from the 2022 Ghana Demographic and Health Survey, linked to 600 georeferenced clusters. A composite cariogenic feeding index was built from bottle feeding, sugar-sweetened beverage consumption, and other cariogenic foods; a child was classified as exposed if any one component was present. Complex survey design was handled by Taylor series linearisation. Determinants were examined using survey-weighted and two-level random-intercept logistic regression, and spatial clustering was assessed using global and local Moran’s I and Getis-Ord Gi* from a five-nearest-neighbour weights matrix.

**Results:** Weighted national prevalence was 52.97% (95% confidence interval 50.19-55.75), ranging from 27.95% in Savannah Region to 73.55% in Western Region, and patterned by residence (63.07% urban versus 43.90% rural) and wealth (32.87% poorest versus 74.45% richest quintile). Wealth and maternal education were the strongest multilevel correlates: the richest households showed roughly four times the odds of exposure relative to the poorest (adjusted odds ratio 4.00), and higher maternal education showed roughly two and a half times the odds relative to no education (adjusted odds ratio 2.51). The intraclass correlation coefficient was 0.219. Global Moran’s I was 0.231 (z = 9.73, p < 0.001), confirming significant positive spatial autocorrelation, with high-high clusters along the southern coastal belt (Western, Central, Greater Accra, Volta) and low-low clusters in the middle and northern belts (Bono, Ahafo, Upper West, Northern, North East).

**Conclusions:** Cariogenic feeding among Ghanaian infants is common, socioeconomically patterned, and spatially clustered rather than random. Oral health promotion through routine growth monitoring and immunisation platforms should be prioritised in the high-high clusters, while closing socioeconomic gradients would yield collateral benefit nationally.

**Trial registration:** Not applicable.

## 1.0 Background

Early childhood caries is defined by the presence of one or more decayed, missing, or filled primary tooth surfaces in a child younger than six years, and it continues to rank among the most common chronic diseases of childhood worldwide, disproportionately affecting children in sub-Saharan Africa where access to preventive and restorative dental care remains limited. Unlike many chronic paediatric conditions, the biological pathway to early childhood caries is comparatively well understood: prolonged and frequent exposure of the primary dentition to fermentable carbohydrates, whether delivered through bottle feeding with sweetened liquids, sugar-sweetened beverages, or energy-dense processed foods, lowers plaque pH and creates the substrate on which cariogenic bacteria, principally mutans streptococci, proliferate. Because these exposures are typically established during infancy, often before the eruption of the full primary dentition, the period from six to twenty-three months of life represents a critical and modifiable window for caries prevention, one that coincides precisely with the period the Demographic and Health Survey programme uses to assess infant and young child feeding.

The nutritional dimensions of infant and young child feeding in Ghana and comparable West African settings have received sustained research attention, with successive analyses of the Ghana Demographic and Health Survey and the Ghana Multiple Indicator Cluster Survey documenting the prevalence of minimum dietary diversity, minimum acceptable diet, and their social and demographic correlates, and identifying maternal education, household wealth, and place of residence as recurring determinants of adequate complementary feeding. [1–4] A parallel body of work, extending across Ghana and several other sub-Saharan African countries, has begun to separate the question of dietary adequacy from that of dietary harm, showing that the same complementary feeding period during which many children fail to meet minimum diversity thresholds is also the period during which sugar-sweetened beverages, sugary snacks, and bottle feeding with sweetened liquids are introduced, sometimes as a substitute for, rather than a complement to, more nutritious foods. [5–9] These cariogenic and obesogenic exposures have been linked not only to early childhood caries but, over a longer horizon, to elevated risk of overweight and early markers of metabolic disease, a concern that has motivated global surveillance of sugar-sweetened beverage intake among children in low- and middle-income countries. [10,11]

What this literature has not yet done, in the Ghanaian setting or in most comparable settings, is treat cariogenic feeding as a single measurable exposure and ask where, geographically, it concentrates. The broader nutrition and health geography literature in West and East Africa has repeatedly shown that undernutrition indicators such as stunting and wasting are not distributed at random across a country’s territory, but instead form statistically identifiable hot spots and cold spots once geographic coordinates are linked to survey clusters and analysed using local indicators of spatial association.[12–19] These studies, conducted for Ethiopia, Uganda, Bangladesh, and Ghana itself, have consistently found that ignoring spatial structure in survey data risks masking substantively important local variation that a purely regional or national tabulation would average away, and that the resulting hot spot maps carry direct programmatic value because they identify the specific districts or clusters where an intervention would have the greatest marginal effect. Cariogenic feeding, despite being conceptually and behaviourally close to the complementary feeding indicators that this literature has already mapped, has not to our knowledge been examined through this same spatial lens using nationally representative Ghanaian data, and the geographic distribution of oral-health-relevant feeding exposures among Ghanaian infants therefore remains, in effect, invisible to policy.

This gap matters for three interconnected reasons. First, Ghana’s oral health workforce and preventive infrastructure are concentrated in a small number of urban centres, so that a policy response calibrated only to a national or regional average prevalence risks either under-resourcing the specific clusters where exposure is most concentrated or spreading limited resources across areas where exposure is already comparatively low. Second, because cariogenic feeding shares upstream determinants, namely household wealth, maternal education, and urban residence, with the wider nutrition transition already documented in Ghana, a spatial pattern that mirrors known socioeconomic gradients would strengthen the case for integrating oral health messaging into existing maternal and child health platforms operating in those same high-burden areas, whereas a spatial pattern that diverges from those gradients would suggest the operation of local factors, such as feeding norms or the marketing reach of sweetened products, that a purely socioeconomic model would miss. Third, from a methodological standpoint, most Ghanaian analyses of infant feeding to date have relied on multilevel or standard logistic regression alone; pairing such models with formal tests of spatial autocorrelation, namely global and local Moran’s I and the Getis-Ord Gi* statistic, provides a check on whether the residual clustering captured by a random cluster intercept has an interpretable geographic signature, information that a variance component alone cannot supply.

Guided by this gap, the present study pursues four specific objectives. First, we estimate the national and subnational prevalence of a composite cariogenic feeding index among Ghanaian children aged 6 to 23 months, using nationally representative data from the 2022 Ghana Demographic and Health Survey. Second, we identify the individual, household, and community-level correlates of this exposure using both survey-weighted and multilevel logistic regression, and we partition the total variance in exposure into its within-cluster and between-cluster components. Third, we test whether cariogenic feeding prevalence is spatially clustered across Ghana’s enumeration areas, using global Moran’s I, and, if clustering is present, we map its local expression using Local Indicators of Spatial Association and the Getis-Ord Gi* statistic to delineate statistically significant hot spots and cold spots. Fourth, we interpret the resulting spatial pattern against the socioeconomic and residential correlates identified in the regression models, in order to assess whether geographic targeting of oral health interventions would add discriminating value beyond what a socioeconomic targeting rule alone would achieve. In addressing these objectives we aim to provide the first spatially explicit, nationally representative account of cariogenic feeding practices in Ghana, and to furnish planners with cluster-level evidence that can inform the geographic prioritisation of infant oral health promotion within existing child health platforms.

## 2.0 Methods

### 2.1 Study design, setting, and data source

This study is a secondary, cross-sectional analysis of the 2022 Ghana Demographic and Health Survey, a nationally representative household survey implemented by the Ghana Statistical Service with technical support from the DHS Program. The survey used a stratified two-stage cluster sampling design in which enumeration areas, serving as primary sampling units, were selected with probability proportional to size within each of the sixteen administrative regions and within the urban and rural strata of each region, followed by systematic selection of households within each selected enumeration area. We used two linked components of the public-use dataset, namely the Children’s Recode, which contains individual-level data for all children born to interviewed women in the five years preceding the survey, and the Geographic Data file, which provides displaced global positioning system coordinates for each of the 618 sampled enumeration clusters.

### 2.2 Analytic sample

The analytic sample was restricted to children aged 6 to 23 completed months at the time of interview, since this is the age band over which the survey’s infant and young child feeding module elicits twenty-four-hour dietary recall, and over which primary tooth eruption is most actively under way. Restricting the sample in this manner is standard practice in the infant and young child feeding literature and yielded an analytic sample of 3,007 children out of 9,353 records in the Children’s Recode.

### 2.3 Construction of the cariogenic feeding index

We defined a binary composite cariogenic feeding index, denoted *CFI*_i_ for child *i*, from three underlying binary indicators, each coded 1 if the exposure was reported and 0 otherwise, with missing responses conservatively coded as unexposed. The first component, bottle feeding (*B*_i_), was derived from the survey item on whether the child was drinking from a bottle with a nipple.

The second component, sweetened beverage consumption (*S*_i_), was derived as the logical union of three items capturing consumption, in the twenty-four hours preceding the interview, of sugary or fizzy drinks, of infant formula, and of other sweetened milk-based or tea-based drinks.

The third component, unhealthy food consumption (*U*_i_), was derived as the logical union of two items capturing consumption of sugary foods such as chocolates, sweets, candies, pastries, cakes, or biscuits, and of other locally defined energy-dense, nutrient-poor foods. Formally,

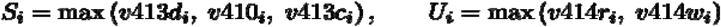

and the composite index was then defined as

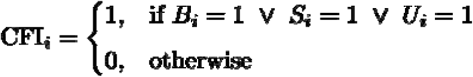

so that a child was classified as exposed to cariogenic feeding if at least one of the three underlying behaviours was reported. This union-based construction follows the logic of composite exposure indices used elsewhere in the infant feeding literature and was chosen in preference to a summed severity score because our primary interest lay in the presence or absence of any cariogenic exposure and in its geographic distribution, rather than in dose-response gradation across the three components.

### 2.4 Covariates

Individual and household covariates considered were child’s age in completed months, child’s sex, mother’s highest educational attainment (no education, primary, secondary, higher), household wealth quintile (poorest to richest, derived by DHS from a household asset index), and place of residence (urban, rural). Region of residence, corresponding to the sixteen 2022 administrative regions, was retained for descriptive stratification.

### 2.5 Accounting for the complex survey design

All national and subnational prevalence estimates and all survey-weighted regression models incorporated the survey’s stratified, clustered design and sampling weights. We defined the survey design object with primary sampling unit identifiers nested within sampling strata, and normalised sampling weights *w*_i_ = *v005*_i_ / 1,000,000 as instructed in the DHS recode manual.

Standard errors were obtained through Taylor series linearisation rather than through the naive assumption of independent and identically distributed observations. For a domain mean such as the weighted CFI prevalence, the point estimate is

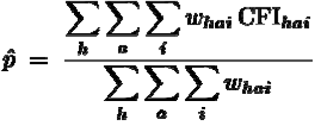

where *h* indexes sampling strata, *a* indexes primary sampling units nested within stratum *h*, and *i* indexes children within cluster *a*, and its linearised variance accounts for the between-cluster component of variability arising from the two-stage design. We additionally computed the design effect,

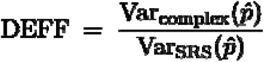

as a diagnostic for the degree of clustering inflation relative to a hypothetical simple random sample of equal size.

### 2.6 Regression models

Two complementary regression strategies were used to characterise the individual, household, and community correlates of cariogenic feeding, each addressing a different source of dependence in the data.

#### Survey-weighted logistic regression

We fitted a survey-weighted generalised linear model with a logit link and quasibinomial error family, which is the standard approach for binary outcomes under complex survey designs because it permits an overdispersion parameter to be estimated rather than fixed at one. For child *i* with covariate vector **x**_i_,

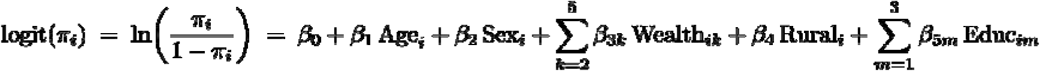

where π_i_ = Pr(*CFI*_i_ = 1 | **x**_i_), wealth and maternal education are entered as sets of indicator variables relative to a reference category (poorest quintile and no education, respectively), and coefficients were exponentiated to yield adjusted odds ratios with Wald-type 95% confidence intervals derived from the design-based standard errors.

#### Two-level random-intercept logistic regression

To decompose the total variance in cariogenic feeding into within-cluster and between-cluster components, and to obtain cluster-specific shrinkage estimates unavailable from a fixed-effects survey model, we additionally fitted a generalised linear mixed model with a random intercept for enumeration cluster, estimated by maximum likelihood using the Laplace approximation. For child *i* nested within cluster *j*,

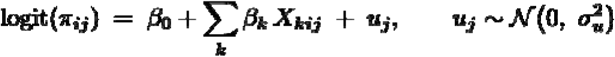

where *u*_j_ is the cluster-level random effect, assumed normally distributed with variance σ²_u_, and *X*_kij_ collects the same fixed covariates used in the survey-weighted model. We first fitted an empty, covariate-free model to obtain the unconditional between-cluster variance, from which the intraclass correlation coefficient, expressing the proportion of total variance in cariogenic feeding attributable to differences between clusters, was computed on the latent logistic scale as

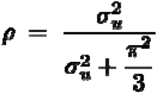

using the standard level 1 variance of π²/3 ≈ 3.29 implied by the logistic distribution. We then fitted the fully adjusted multilevel model with the same covariate set as the survey-weighted model, to allow comparison of point estimates obtained under the two complementary approaches to handling the clustered structure of the data.

### 2.7 Linking survey clusters to geographic coordinates

Cluster-level global positioning system coordinates from the Geographic Data file were merged to the Children’s Recode by the shared enumeration cluster identifier, after excluding clusters with missing or displaced-to-zero coordinates, a standard data quality step in DHS spatial analysis. For each of the resulting geo-referenced clusters we computed the cluster-level cariogenic feeding prevalence,

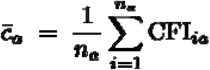

where *n*_a_ is the number of sampled children aged 6 to 23 months in cluster *a*. Of the 618 clusters in the geographic dataset, 600 had both valid coordinates and at least one eligible child and constituted the spatial analytic unit for all subsequent geostatistical procedures.

### 2.8 Spatial weights matrix

Spatial relationships among the 600 clusters were represented using a *k*-nearest-neighbour contiguity structure with *k* = 5, chosen to ensure a fully connected graph with a fixed number of neighbours per cluster regardless of local cluster density, which varies substantially between the densely sampled south and the sparsely sampled north of Ghana. Neighbour sets *N*_k_(*a*) were computed from the projected geographic coordinates, and the resulting binary neighbour matrix was row-standardised so that each cluster’s neighbour weights summed to one,

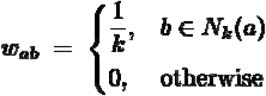

### 2.9 Global spatial autocorrelation

Global spatial clustering of cluster-level cariogenic feeding prevalence was tested using Moran’s I statistic,

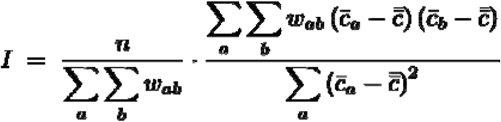

where *n* (the overall mean cluster prevalence) is the unweighted mean of cluster-level prevalence across all clusters, and *w*_ab_ are the row-standardised spatial weights defined above. Under the randomisation null hypothesis of no spatial pattern, the statistic has expectation

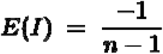

and a variance obtained from the standard randomisation formula that accounts for the kurtosis of the observed prevalence distribution, from which a standardised test statistic

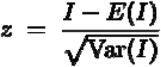

was compared against the standard normal distribution to obtain a one-sided p-value against the alternative hypothesis of positive spatial autocorrelation, that is, that geographically proximate clusters have more similar prevalence than would be expected under spatial randomness.

### 2.10 Local indicators of spatial association

To localise the global pattern, we computed the Local Moran’s I statistic for each cluster *a*,

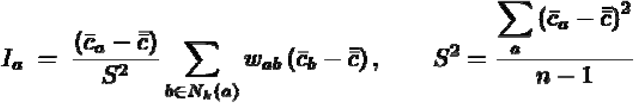

with conditional randomisation used to assess statistical significance of each local statistic, and clusters classified into five mutually exclusive categories at the α = 0.05 threshold according to the sign of the cluster’s own deviation from the mean and the sign of its spatially lagged deviation: high-high, indicating a cluster with above-average prevalence surrounded by other above-average clusters; low-low, indicating a below-average cluster surrounded by other below-average clusters; high-low and low-high, indicating spatial outliers; and not significant. We further computed the Getis-Ord Gi* statistic to identify hot spots and cold spots of high or low prevalence irrespective of the sign classification used by Local Moran’s I,

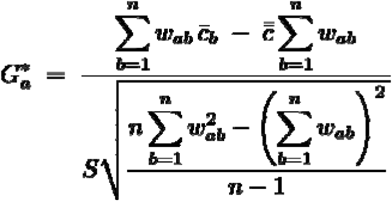

where *S* is the standard deviation of cluster-level prevalence across all *n* clusters. Positive, large- magnitude G_a_* values identify statistically significant hot spots, that is, clusters whose value and whose neighbours’ values are consistently higher than the global mean, while negative, large-magnitude values identify cold spots.

### 2.11 Software and reproducibility

All analyses were conducted in R version 4.5.2. Survey-weighted estimation used the survey package, spatial data handling used sf, spatial weights construction and autocorrelation statistics used spdep, multilevel modelling used lme4, and choropleth and point-pattern mapping used tmap, overlaid on first-level administrative boundaries obtained through the geodata interface to the Database of Global Administrative Areas. A two-sided α of 0.05 was used for all hypothesis tests other than the one-sided global Moran’s I test described above, for which a one-sided alternative was specified a priori because the hypothesis under investigation was positive, not negative, spatial autocorrelation.

### 2.12 Ethical considerations and use of artificial intelligence

This analysis used de-identified, publicly available secondary survey data and did not require separate ethical clearance beyond that already obtained by the DHS Program and the Ghana Statistical Service for the original survey; further detail is provided in the Declarations section. A large language model based writing assistant was used during manuscript preparation to support language editing and formatting of the draft text; the assistant did not generate, select, or interpret any statistical result, and all analytic code, output, and scientific conclusions are the sole responsibility of the human authors, consistent with the journal’s authorship policy on the use of such tools.

## 3.0 Results

### 3.1 Sample characteristics and unadjusted component prevalence

Of the 3,007 children aged 6 to 23 months in the analytic sample, unweighted component prevalence was 19.4% for bottle feeding (582 children), 21.5% for sweetened beverage consumption (647 children), and 24.2% for consumption of other cariogenic foods (728 children), giving an unweighted composite prevalence of 46.5%. Once sampling weights, stratification, and clustering were incorporated, the national weighted prevalence of the composite cariogenic feeding index was 52.97% (standard error 1.42%, 95% confidence interval 50.19–55.75%), appreciably higher than the unweighted figure, a divergence consistent with the differential sampling and non-response weighting applied across strata and indicating that unweighted tabulation alone would have understated national exposure.

### 3.2 Prevalence by region, residence, and wealth

Weighted prevalence differed substantially across Ghana’s sixteen regions (Table 1). Prevalence was highest in the Western Region (73.55%, standard error 3.82) and lowest in the Savannah Region (27.95%, standard error 4.00), a range of nearly forty-six percentage points between the highest and lowest-prevalence regions. Five regions exceeded 60%, namely Western, Central, Greater Accra, Volta, and Eastern, while four regions in the northern and middle belts, namely Savannah, Oti, Northern, and Upper West, remained below 35%.

**Table 1.** Weighted prevalence of cariogenic feeding by administrative region.

| Region | Prevalence (%) | Standard error |
| --- | --- | --- |
| Western | 73.55 | 3.82 |

| **Region** | **Prevalence (%)** | **Standard error** |
| --- | --- | --- |
| Central | 68.20 | 4.09 |
| Greater Accra | 67.47 | 5.29 |
| Volta | 61.75 | 4.57 |
| Eastern | 60.60 | 5.83 |
| Ashanti | 59.01 | 4.03 |
| Upper East | 47.52 | 4.66 |
| Ahafo | 49.16 | 5.64 |
| Western North | 46.11 | 7.27 |
| Bono East | 44.09 | 4.67 |
| Bono | 39.25 | 4.53 |
| North East | 37.92 | 4.39 |
| Upper West | 33.09 | 5.16 |
| Northern | 31.45 | 3.39 |
| Oti | 29.57 | 4.47 |
| Savannah | 27.95 | 4.00 |

*2022 Ghana Demographic and Health Survey, children aged 6–23 months (n = 3,007). Estimates are weighted for the survey's stratified, clustered sample design; standard errors were derived by Taylor series linearisation. Regions are ordered from highest to lowest prevalence.*

Cariogenic feeding was more common among urban children (63.07%, standard error 1.99) than rural children (43.90%, standard error 1.92), a gap of just over nineteen percentage points. A pronounced and monotonic wealth gradient was also evident (Table 2), rising from 32.87% in the poorest quintile to 74.45% in the richest, a more than two-fold increase across the wealth distribution.

**Table 2.** Weighted prevalence of cariogenic feeding by household wealth quintile.

| Wealth quintile | Prevalence (%) | Standard error |
| --- | --- | --- |
| Poorest | 32.87 | 3.01 |
| Poorer | 41.67 | 2.29 |
| Middle | 58.20 | 2.53 |
| Richer | 66.75 | 3.17 |
| Richest | 74.45 | 3.49 |

5 *Wealth quintile is the standard Demographic and Health Survey household asset-based index; estimates are survey-weighted with Taylor series linearised standard errors.*

### 3.3 Correlates of cariogenic feeding: survey-weighted logistic regression

In the survey-weighted logistic regression model (Table 3), older child age was associated with higher odds of cariogenic feeding (adjusted odds ratio 1.04 per completed month, 95% confidence interval 1.02–1.06, p < 0.001), consistent with cumulative exposure as complementary feeding progresses. Child sex was not associated with exposure. Household wealth showed a strong, graded association: relative to the poorest quintile, the odds of cariogenic feeding were more than doubled in the middle quintile (adjusted odds ratio 2.10, 95% confidence interval 1.44-3.06), nearly tripled in the richer quintile (adjusted odds ratio 2.72, 95% confidence interval 1.74-4.25), and approximately tripled in the richest quintile (adjusted odds ratio 2.99, 95% confidence interval 1.72-5.19). Urban-rural residence was not independently associated with exposure once wealth and education were held constant, indicating that the unadjusted urban excess described above operates substantially through the socioeconomic composition of urban households rather than through residence per se. Maternal education showed the steepest gradient of any covariate: relative to children of mothers with no education, the adjusted odds of cariogenic feeding were 43% higher among children of mothers with primary education, more than double among those with secondary education (adjusted odds ratio 2.21, 95% confidence interval 1.61-3.02), and nearly five-fold among those whose mothers had higher education (adjusted odds ratio 4.72, 95% confidence interval 2.84-7.85).

**Table 3.** Survey-weighted logistic regression of cariogenic feeding on individual and household covariates.

| **Covariate** | **Adjusted OR** | **95% CI** | **p-value** |
| --- | --- | --- | --- |
| Child age (months) | 1.04 | 1.02–1.06 | < 0.001 |
| Female sex (ref. male) | 1.02 | 0.83–1.25 | 0.853 |
| Wealth: poorer (ref. poorest) | 1.26 | 0.90–1.77 | 0.182 |
| Wealth: middle | 2.10 | 1.44–3.06 | < 0.001 |
| Wealth: richer | 2.72 | 1.74–4.25 | < 0.001 |
| Wealth: richest | 2.99 | 1.72–5.19 | < 0.001 |
| Rural residence (ref. urban) | 0.92 | 0.71–1.20 | 0.541 |
| Maternal education: primary (ref. none) | 1.43 | 1.01–2.02 | 0.042 |
| Maternal education: secondary | 2.21 | 1.61–3.02 | < 0.001 |

| Covariate | Adjusted OR | 95% CI | p-value |
| --- | --- | --- | --- |
| Maternal education: higher | 4.72 | 2.84–7.85 | < 0.001 |

*Model estimated by survey-weighted quasibinomial logistic regression accounting for stratification, clustering, and sampling weights; reference categories are poorest wealth quintile, urban residence, and no maternal education. OR: odds ratio; CI: confidence interval.*

### 3.4 Multilevel decomposition of variance and cluster-adjusted correlates

The empty, covariate-free multilevel model estimated a between-cluster variance of σ ²_u_ = 0.469 on the logistic scale, yielding an intraclass correlation coefficient of ρ = 0.219: approximately twenty-two percent of the total variability in cariogenic feeding among Ghanaian children aged 6 to 23 months lay between enumeration clusters rather than between individual children within the same cluster, a magnitude sufficiently large to justify both the multilevel modelling strategy adopted here and the subsequent formal test for spatial, as opposed to merely hierarchical, structure in that between-cluster variance.

The fully adjusted multilevel model reproduced the direction and approximate magnitude of every association identified in the survey-weighted model (Table 4), lending consistency to the substantive conclusions across two analytically distinct approaches to handling clustering. Household wealth remained the covariate with the largest effect size, with children in the richest households having approximately four times the odds of cariogenic feeding relative to the poorest (adjusted odds ratio 4.00, 95% confidence interval derived from a standard error of 0.203 on the log-odds scale), and maternal higher education remained associated with more than double the odds relative to no education (adjusted odds ratio 2.51). Rural residence was associated with modestly, though not statistically significantly, lower odds after accounting for the cluster random intercept (adjusted odds ratio 0.89, p = 0.315), again suggesting that the socioeconomic composition of clusters, rather than urban-rural status as such, is the more proximal determinant.

**Table 4.**
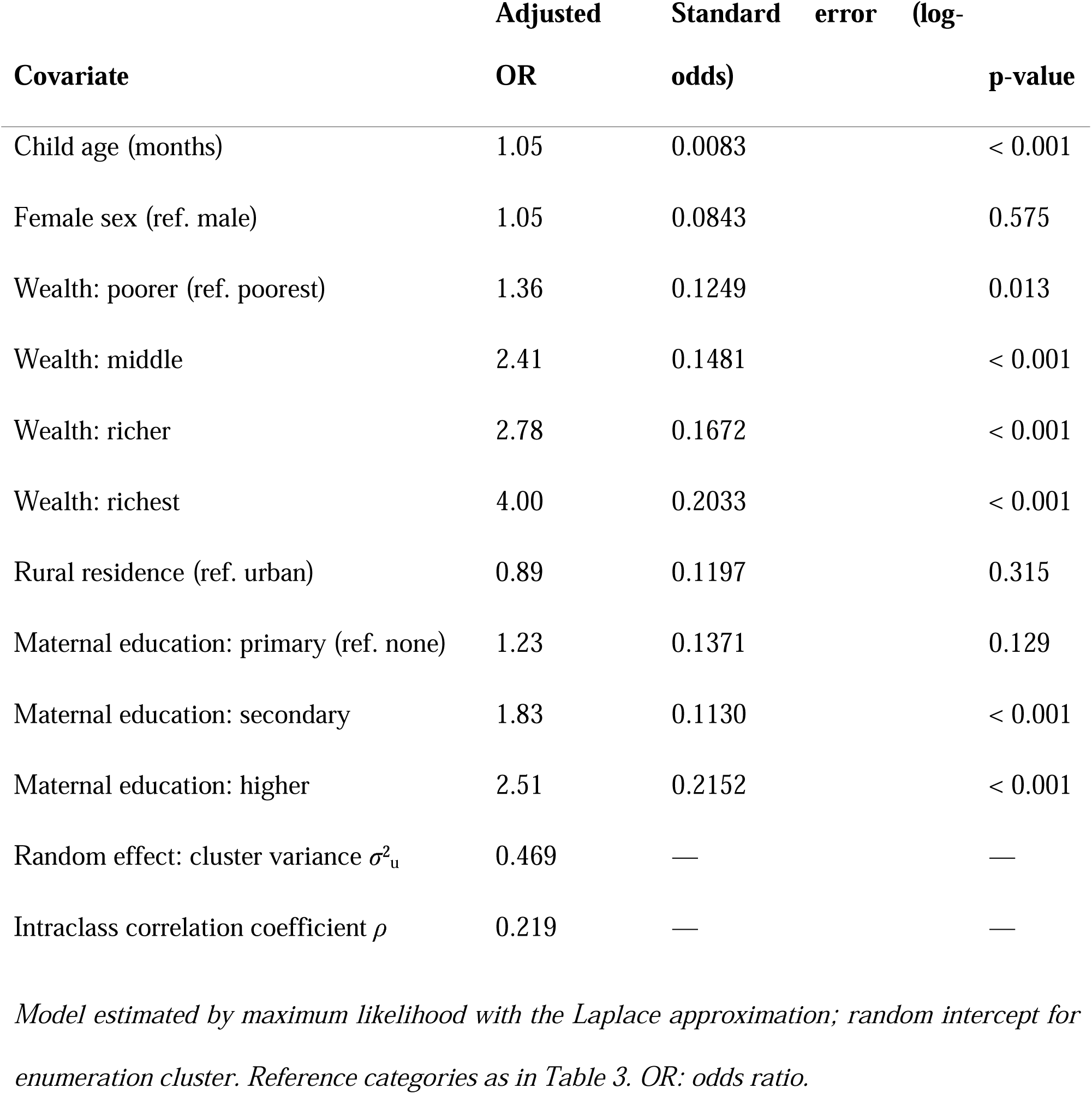
Two-level random-intercept logistic regression of cariogenic feeding, children nested within enumeration clusters.

| Covariate | Adjusted OR | Standard error (log-odds) | p-value |
| --- | --- | --- | --- |
| Child age (months) | 1.05 | 0.0083 | < 0.001 |
| Female sex (ref. male) | 1.05 | 0.0843 | 0.575 |
| Wealth: poorer (ref. poorest) | 1.36 | 0.1249 | 0.013 |
| Wealth: middle | 2.41 | 0.1481 | < 0.001 |
| Wealth: richer | 2.78 | 0.1672 | < 0.001 |
| Wealth: richest | 4.00 | 0.2033 | < 0.001 |
| Rural residence (ref. urban) | 0.89 | 0.1197 | 0.315 |
| Maternal education: primary (ref. none) | 1.23 | 0.1371 | 0.129 |
| Maternal education: secondary | 1.83 | 0.1130 | < 0.001 |
| Maternal education: higher | 2.51 | 0.2152 | < 0.001 |
| Random effect: cluster variance $\sigma^2_u$ | 0.469 | — | — |
| Intraclass correlation coefficient $\rho$ | 0.219 | — | — |

### 3.5 Global spatial autocorrelation

Among the 600 clusters with valid geographic coordinates and eligible children, cluster-level cariogenic feeding prevalence ranged from 0% to 100%, with a mean of approximately 51% and visibly non-random spatial arrangement on inspection of the choropleth (Fig. 1). Global Moran’s I confirmed this impression statistically: *I* = 0.231 against a randomisation expectation of *E*(*I*) = - 0.0017 and variance 0.00057, giving a standardised test statistic of *z* = 9.73 and a p-value below 2.2 × 10^-16^. This result provides strong evidence against the null hypothesis of spatial randomness and in favour of positive spatial autocorrelation: clusters located near one another tend to report more similar cariogenic feeding prevalence than would be expected by chance, a pattern consistent with shared local determinants such as market access to sweetened products, area-level socioeconomic composition, or locally transmitted infant feeding norms operating at a spatial scale coarser than the individual household.

**Figure 1.**
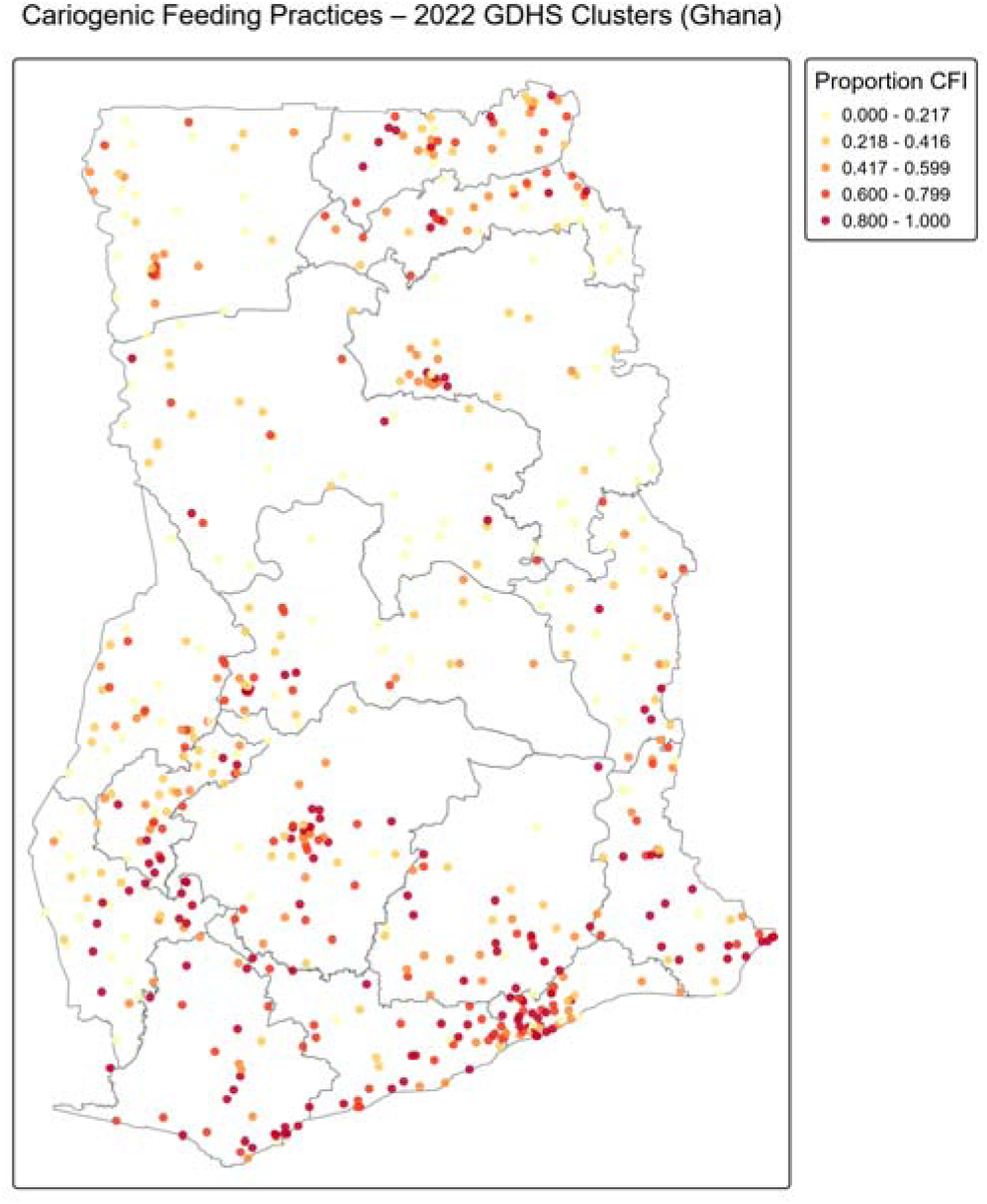
Cluster-level cariogenic feeding prevalence, Ghana, 2022. Choropleth of weighted prevalence across 600 georeferenced Demographic and Health Survey enumeration clusters, overlaid on first-level administrative boundaries. Colour intensity denotes quintile of cluster- level prevalence, from lowest (pale yellow) to highest (dark red).

### 3.6 Local clustering: hot spots, cold spots, and LISA quadrants

The Getis-Ord Gi* statistic (Fig. 2) delineated a coherent hot spot of elevated cariogenic feeding running along the southern coastal and mid-southern belt of the country, most prominently in a dense band spanning the Greater Accra coastline, the southern Volta and Eastern regions, and a compact, high-magnitude cluster in central Ashanti in the vicinity of the Kumasi metropolitan area, together with a further concentration along the far southwestern coast of the Western Region. Conversely, a broad cold spot of significantly negative Gi* values extended across the entire northern half of the country, encompassing Upper West, most of the Northern Region, North East, and parts of Bono and Ahafo in the transitional middle belt, indicating clusters whose cariogenic feeding prevalence, together with that of their neighbours, was consistently lower than the national mean.

**Figure 2.**
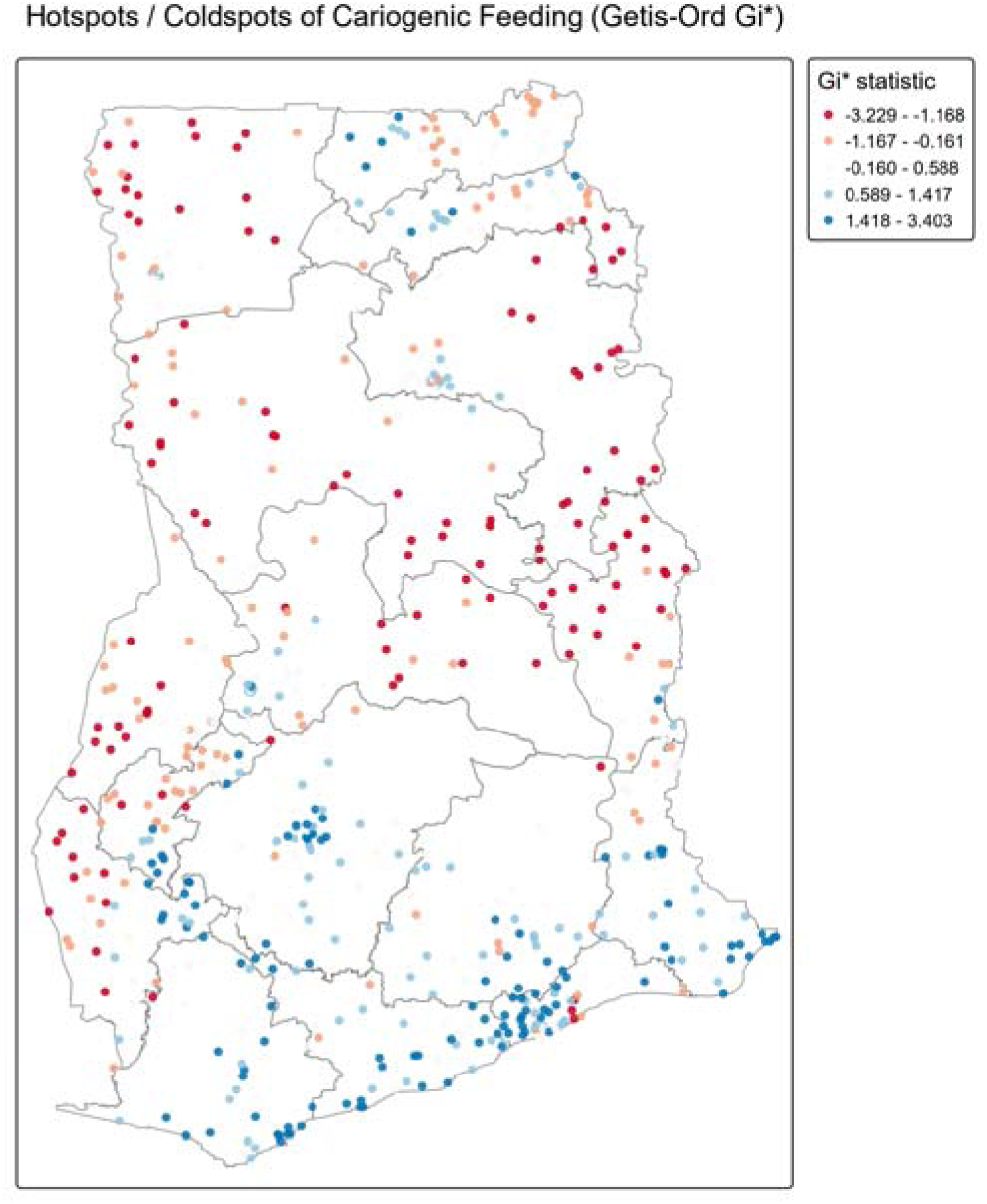
Hot spot and cold spot map of cariogenic feeding. Getis-Ord Gi* statistic computed from a five-nearest-neighbour spatial weights matrix. Blue denotes statistically significant hot spots of high prevalence; red denotes significant cold spots of low prevalence.

The Local Moran’s I classification (Fig. 3, Table 5) corroborated this pattern while adding the outlier categories that the Gi* statistic does not distinguish. High-high clusters, indicating a statistically significant concentration of high-prevalence clusters surrounded by other high-prevalence clusters, were most heavily concentrated in the Greater Accra coastal fringe, in a compact peri-urban Kumasi cluster within Ashanti, and along the southwestern coast of the Western Region. Low-low clusters, indicating a significant concentration of low-prevalence clusters surrounded by other low-prevalence clusters, were dispersed across Upper West, Northern, North East, and the Bono-Ahafo-Ashanti middle belt. A small number of high-low and low-high spatial outliers, representing clusters whose prevalence diverged sharply from their immediate neighbourhood, were scattered mainly along the boundary between the high-prevalence south and the low-prevalence north, consistent with a genuine spatial transition zone rather than isolated data anomalies. The large majority of clusters did not reach the conventional significance threshold for local clustering, which is the expected pattern when a moderate, though highly significant, global autocorrelation coefficient is decomposed into its local contributions, since global clustering of the observed magnitude is typically carried by a comparatively small number of strongly clustered locations rather than by uniform local significance across the entire map.

**Figure 3.**
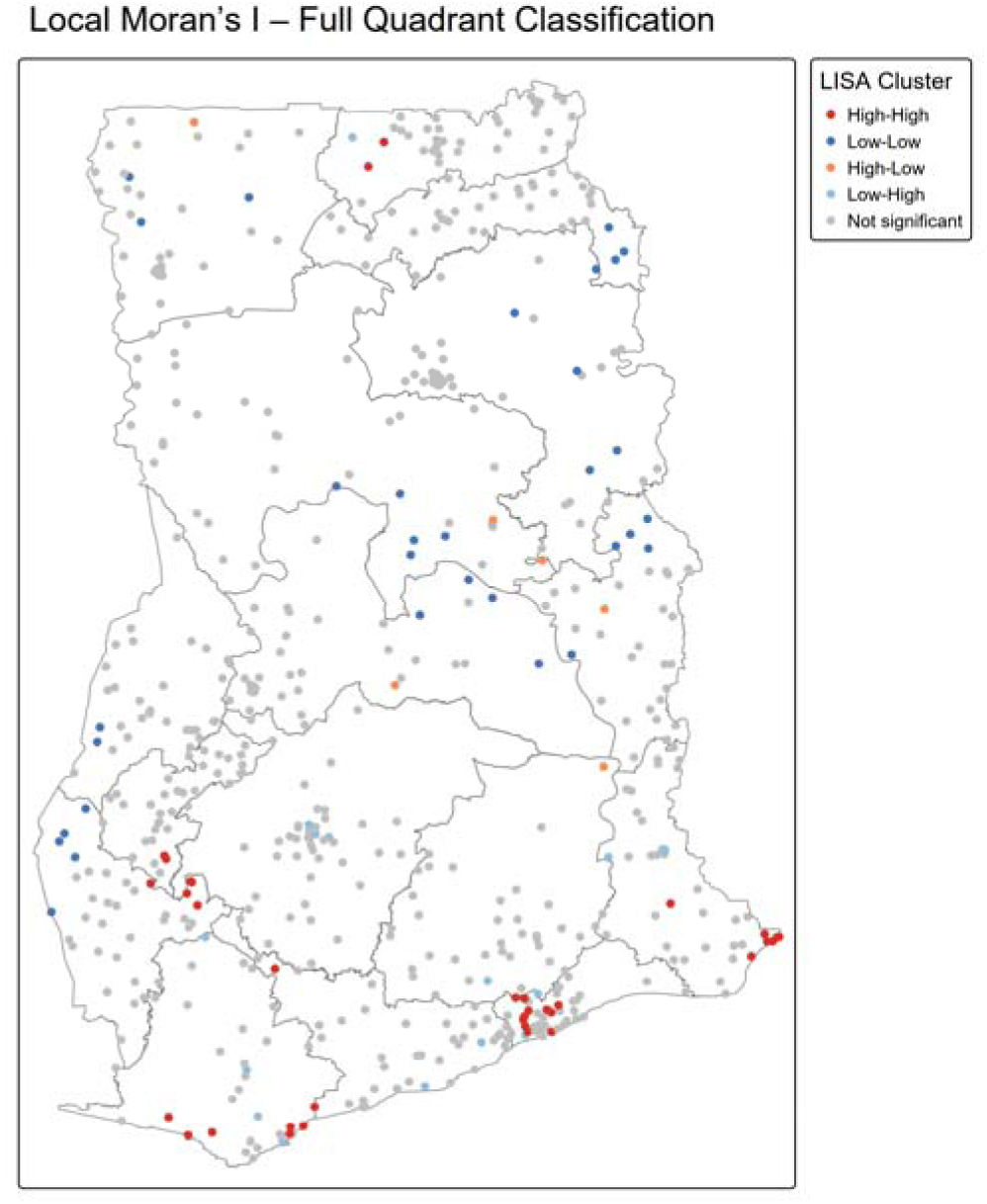
Local Moran’s I cluster and outlier classification. High-high, low-low, high-low, and low-high categories of cariogenic feeding prevalence, classified at the α *= 0.05 significance threshold; grey denotes clusters with no significant local association*.

**Table 5.** Summary of Local Indicators of Spatial Association (LISA) for cluster-level cariogenic feeding prevalence.

| LISA category | Geographic concentration |
| --- | --- |
| High-high | Greater Accra coastal fringe; peri-urban Kumasi (Ashanti); southwestern Western Region coast |
| Low-low | Upper West; Northern; North East; Bono-Ahafo-Ashanti middle belt |
| High-low / Low-high (spatial outliers) | Scattered along the south-north transition zone |
| Not significant | Majority of clusters nationally, distributed across all regions |

## 4.0 Discussion

The central finding of this study is that cariogenic feeding among Ghanaian infants and young children is not merely common, affecting roughly one in two children nationally, but that its distribution across the country’s territory is patterned in a manner that a conventional regional or wealth-stratified tabulation only partly captures. Global Moran’s I of 0.231 is, in absolute terms, a moderate coefficient, yet the associated z-statistic of 9.73 places the observed clustering many standard deviations beyond what spatial randomness would produce, a strength of signal broadly comparable to that reported for undernutrition indicators in neighbouring Ethiopia and in Bangladesh, where local indicators of spatial association have likewise identified compact hot spots embedded within a largely non-significant national canvas.[12,13,17–19] That comparable clustering strength has now been demonstrated for a caries-relevant feeding exposure, rather than only for undernutrition, suggests that the geographic logic long applied to Ghana’s nutrition transition extends naturally to its emerging oral health transition, and that the two phenomena may in places share a common spatial fingerprint even though they are conceptually distinct outcomes.

The socioeconomic gradient identified here, rising from roughly one in three children exposed in the poorest quintile to roughly three in four exposed in the richest, situates cariogenic feeding as a marker of relative affluence rather than of deprivation, an orientation that runs counter to the pattern typically reported for undernutrition and stunting in the same population but that aligns closely with what has been documented for sugar-sweetened beverage and processed snack consumption elsewhere on the continent and beyond.[7–10,20] This inversion carries a specific implication for programme design: interventions built on Ghana’s existing poverty-targeting infrastructure, which was designed with undernutrition and food insecurity in mind, will not automatically reach the households at greatest risk of cariogenic feeding, and a purely wealth-based targeting rule may in fact point resources away from, rather than toward, the areas of highest exposure. The persistence of a strong maternal education gradient, with children of mothers who attained higher education exposed at roughly two and a half to nearly five times the odds of children of mothers with no education depending on the modelling approach, further suggests that exposure to sweetened products and formula-fed bottle feeding in Ghana still carries some of the aspirational, modernity-linked connotation that has been described in earlier qualitative and survey-based accounts of infant feeding transitions in West Africa, in which formula, sweetened beverages, and packaged snack foods are perceived by better-resourced, better-educated caregivers as convenient or even nutritionally superior alternatives to traditional weaning foods.[5,6,9] Read against the region-level findings, in which the wealthier, more urbanised south of the country carries substantially higher prevalence than the poorer, more rural north, the spatial and socioeconomic evidence reinforce one another, and this convergence is itself informative: it indicates that the observed hot spots in Greater Accra, coastal Western Region, and peri-urban Kumasi are not an artefact of unmeasured local factors operating independently of socioeconomic status, but rather the geographic expression of a socioeconomic and educational gradient that happens to be spatially concentrated in the country’s most urbanised corridor.

At the same time, the finding that urban-rural residence lost statistical significance once wealth and maternal education were accounted for, in both the survey-weighted and the multilevel model, indicates that residence functions largely as a proxy for household resources and market access rather than as an independent causal pathway, a distinction with practical consequence for how “urban” risk should be operationalised in future surveillance: crude urban-rural stratification, still the most commonly reported disaggregation in Ghanaian nutrition surveillance, is likely to understate the true concentration of cariogenic feeding risk within the wealthier, more educated subset of urban households, and a finer-grained wealth-by-residence or spatially explicit stratification would better serve programme targeting than residence alone. The intraclass correlation coefficient of 0.219 obtained from the empty multilevel model is itself a substantively important result independent of the spatial analysis that followed it, because it quantifies, on a standard and interpretable scale, how much of the variation in this exposure is a property of place rather than of the individual child or household; a value in this range is broadly consistent with cluster-level variance components reported for other behavioural and nutritional exposures in multilevel analyses of Demographic and Health Survey data across the region, and it provided the a priori justification for testing whether that between-cluster variance carried spatial structure, which the subsequent Moran’s I and Getis–Ord Gi* analyses confirmed that it did.

From an oral health systems perspective, the practical value of the local, as opposed to only the global or regional, analysis lies in its capacity to discriminate within regions that a regional tabulation alone would treat as internally homogeneous. Ashanti Region, for example, registered a weighted regional prevalence of 59.0%, a figure that on its own would place it in the upper-middle range of Ghana’s sixteen regions and would not, by itself, identify it as a priority area distinct from several other regions clustered around a similar prevalence. The local analysis, however, isolated a compact, statistically significant high-high cluster within Ashanti, concentrated in and around the Kumasi metropolitan area, embedded within a region that otherwise contains a substantial low-low pocket in its more rural districts; a regional average of 59.0% is, in effect, an arithmetic compromise between these two locally distinct realities, and an intervention allocated at the regional level, without reference to the underlying local pattern, would necessarily under-serve the Kumasi hot spot while over-serving the region’s low-prevalence rural periphery. This is precisely the kind of within-region heterogeneity that the broader African spatial nutrition literature has flagged as a reason to prefer cluster- or district-level targeting over regional averages wherever the underlying survey data permit it, [12,15,16,19] and the present findings extend that methodological argument, for the first time to our knowledge, to a specifically caries-relevant feeding exposure in Ghana.

Several limitations warrant explicit acknowledgement. The cross-sectional design of the Demographic and Health Survey precludes causal inference about the direction or timing of the associations reported here, and in particular cannot establish whether locally clustered cariogenic feeding reflects diffusion of norms through community networks, shared exposure to local food retail environments, unmeasured confounding by area-level infrastructure, or some combination of these mechanisms. The composite cariogenic feeding index, while conceptually grounded in established caries aetiology, necessarily collapses heterogeneous exposures, differing in frequency, quantity, and cariogenic potential, into a single binary indicator, and future work using the full ordinal or frequency information available in the underlying items could usefully test whether the spatial pattern reported here is driven disproportionately by one component, such as bottle feeding, rather than by the composite as a whole. The geographic coordinates supplied with Demographic and Health Survey data are randomly displaced by a small distance for confidentiality, a standard feature of the public-use geographic dataset that introduces a modest degree of positional noise into cluster-level spatial statistics, though this displacement is generally considered to have limited practical effect on cluster-level analyses conducted at the scale used here. Finally, although we linked the observed spatial pattern to the socioeconomic and educational correlates identified in the regression models, we did not have access to area-level covariates such as the density of retail outlets selling sweetened beverages or infant formula, which would be required to formally test, rather than infer, the market-access mechanism proposed above as one plausible driver of the observed hot spots, and this represents a natural direction for subsequent research combining survey and geospatial retail data.

## 5.0 Conclusions

Cariogenic feeding practices affect approximately one in two Ghanaian children aged 6 to 23 months, and both their individual-level determinants and their geographic distribution point toward household wealth, maternal education, and urban, market-connected living environments as the principal correlates of exposure, rather than toward the poverty-linked pathways that dominate Ghana’s undernutrition profile. The demonstration of significant, spatially coherent hot spots along the southern coastal and peri-Kumasi corridor, set against a broad northern and middle-belt cold spot, indicates that cariogenic feeding in Ghana is amenable to geographically targeted preventive action in a way that a national or purely regional prevalence figure could not reveal. We recommend that infant oral health counselling, delivered through existing growth monitoring, immunisation, and antenatal and postnatal care platforms, be prioritised within the identified high-high clusters, with particular attention to caregivers with higher educational attainment who may currently perceive sweetened beverages, formula, and packaged snacks as benign or even advantageous choices, while national efforts to strengthen regulation and labelling of sweetened products marketed for infants and young children would be expected to yield the greatest population-level benefit precisely where wealth, education, and urban market access, and consequently cariogenic feeding, are most concentrated.

## List of abbreviations

CFI: Cariogenic Feeding Index
DHS: Demographic and Health Survey
GDHS: Ghana Demographic and Health Survey
GPS: Global Positioning System
ICC: Intraclass Correlation Coefficient
IYCF: Infant and Young Child Feeding
LISA: Local Indicators of Spatial Association
OR: Odds Ratio
PSU: Primary Sampling Unit
SE: Standard Error.

## Declarations

### Ethics approval and consent to participate

This analysis used publicly available, de-identified secondary data from the Ghana Demographic and Health Surveys, obtained with permission from the DHS Program, ICF International. The original surveys received ethical approval from the Ghana Health Service Ethics Review Committee and the ICF International Institutional Review Board, and all respondents provided informed consent at the time of interview. No additional ethical approval was sought for this secondary analysis, consistent with DHS Program data use policy.

## Consent for publication

Not applicable.

## Availability of data and materials

The dataset used in this study is publicly available from the DHS Program upon registration and approved request at https://dhsprogram.com. The cluster- level cariogenic feeding summary data and derived spatial statistics generated during the current study are available from the corresponding author on reasonable request.

## Authors’ contributions

OAI and AI conceived, designed, analyzed, and wrote major parts of the manuscript. AHS performed data curation, formal analysis, writing, reviewing, and editing. AIS wrote the methodology and discussion and performed data quality checks on the spatial covariates. The final draft of the manuscript was reviewed, edited, and revised for intellectual content, read, and approved by all authors.

## Data Availability

The dataset used in this study is publicly available from the DHS Program upon registration and approved request at https://dhsprogram.com. The cluster-level cariogenic feeding summary data and derived spatial statistics generated during the current study are available from the corresponding author on reasonable request.

## Acknowledgements

The authors thank the Ghana Statistical Service, the Ghana Health Service, and the DHS Program for making the 2022 Ghana Demographic and Health Survey data publicly available for secondary analysis.

## Use of generative artificial intelligence

A generative artificial intelligence-based language model was used solely to assist with language refinement and formatting during manuscript preparation, in accordance with the journal’s policy that such tools do not qualify for authorship.

All data analysis, interpretation of results, and scientific content are the responsibility of the named human authors.

